# Decoding the biology of ethnic differences in asthma

**DOI:** 10.1101/2024.11.08.24316950

**Authors:** Duan Ni, Ralph Nanan

## Abstract

Ethnicity seems to be a risk factor for asthma and its exacerbation, but no general consensus has been reached so far. Ethnic diversity and differences have long been overlooked and most studies in these regards are observational, lacking in-depth mechanistic insights.

Harnessing comprehensive transcriptomic data at organ system and single cell levels, we comparatively analyzed the signalling landscapes of African American/Black (AA) and other ethnicities. In healthy individuals, most AA PBMC subsets exhibited elevated IL2-STAT5 signals, which is critically implicated in asthma pathogenesis. This extended to various compartments (bronchial and airway epithelium, circulating CD4^+^ T cells and blood) in asthma, where AA upregulated IL2-STAT5 pathway relative to other ethnicities. Mechanistically, the increased IL2-STAT5 signalling correlated with enhanced T cell migration and recruitment. Together, they might contribute to the greater predisposition towards asthma exacerbation in AA.

This first-of-its-kind study sheds some in-depth insights into the biology of ethnic differences in asthma. Our findings, if further validated, will be valuable for future asthma research and clinical practice like therapeutic development and treatment guidance, highlighting the need for ethnicity diversity and inclusions in these areas.

---

Ethnicity may impact asthma, with some studies suggesting that African Americans are of higher risk and tend to experience more severe disease symptoms^1,2^. However, there is no general consensus on this issue^3^, as research has historically focused predominantly on white/Caucasian populations. Additionally, the mechanisms underlying possible ethnic differences in asthma remain poorly understood.

In the pathogenesis of asthma, the IL2-STAT5 signalling plays a crucial role^4,5^. Leveraging a published cellular indexing of transcriptomes and epitopes sequencing (CITE-seq) dataset analyzing healthy European American/White (EA) and African American/Black (AA) individuals, we extensively compared the IL2-STAT5 signalling across various PBMC subsets based on their gene set scores. Strikingly, in all T cell and natural killer cell subsets, IL2-STAT5 signalling was elevated in AA. Most AA-derived monocyte subsets, naive and germinal centre B cells also exhibited similar patterns, while memory B cells, CD34^+^ progenitors and dendritic cell subsets showed no difference between AA and EA (Figure1). These data suggests that under basal conditions, AA PBMCs generally upregulate IL2-STAT5 signalling compared with EA.

**Figure 1.**
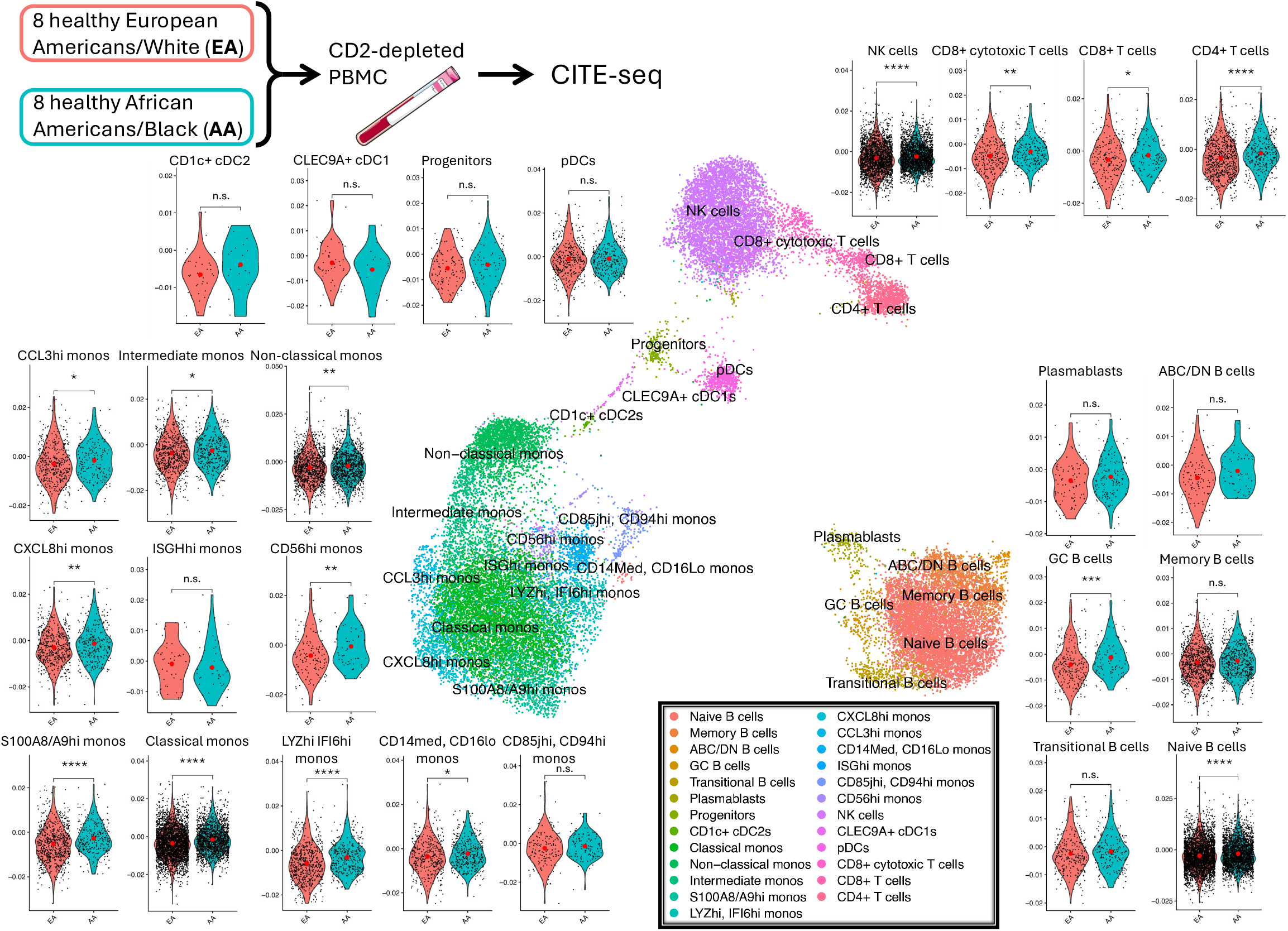
IL2-STAT5 signalling landscape in healthy African American/Black (AA) and European American/White (EA) peripheral blood mononuclear cells (PBMCs). Cellular indexing of transcriptomes and epitopes sequencing (CITE-seq) dataset was retrieved from GSE189050. In healthy controls, gene set scores for IL2-STAT5 signalling were calculated for AA (cyan) and EA (pink), with the red dots representing the average gene set score levels of the corresponding subsets.

We next exhaustively surveyed asthma-related transcriptomic studies available on Gene Expression Omnibus. We found that most of these studies actually did not report or consider the factor of ethnicity/race and only 8 studies reported the ethnicity/race of their participants. Among them, 3 studies were excluded for low data quality and 5 studies (4 cross-sectional and 1 longitudinal studies) were used for further comparative analyses (Figure2A, Supplementary Information).

**Figure 2.**
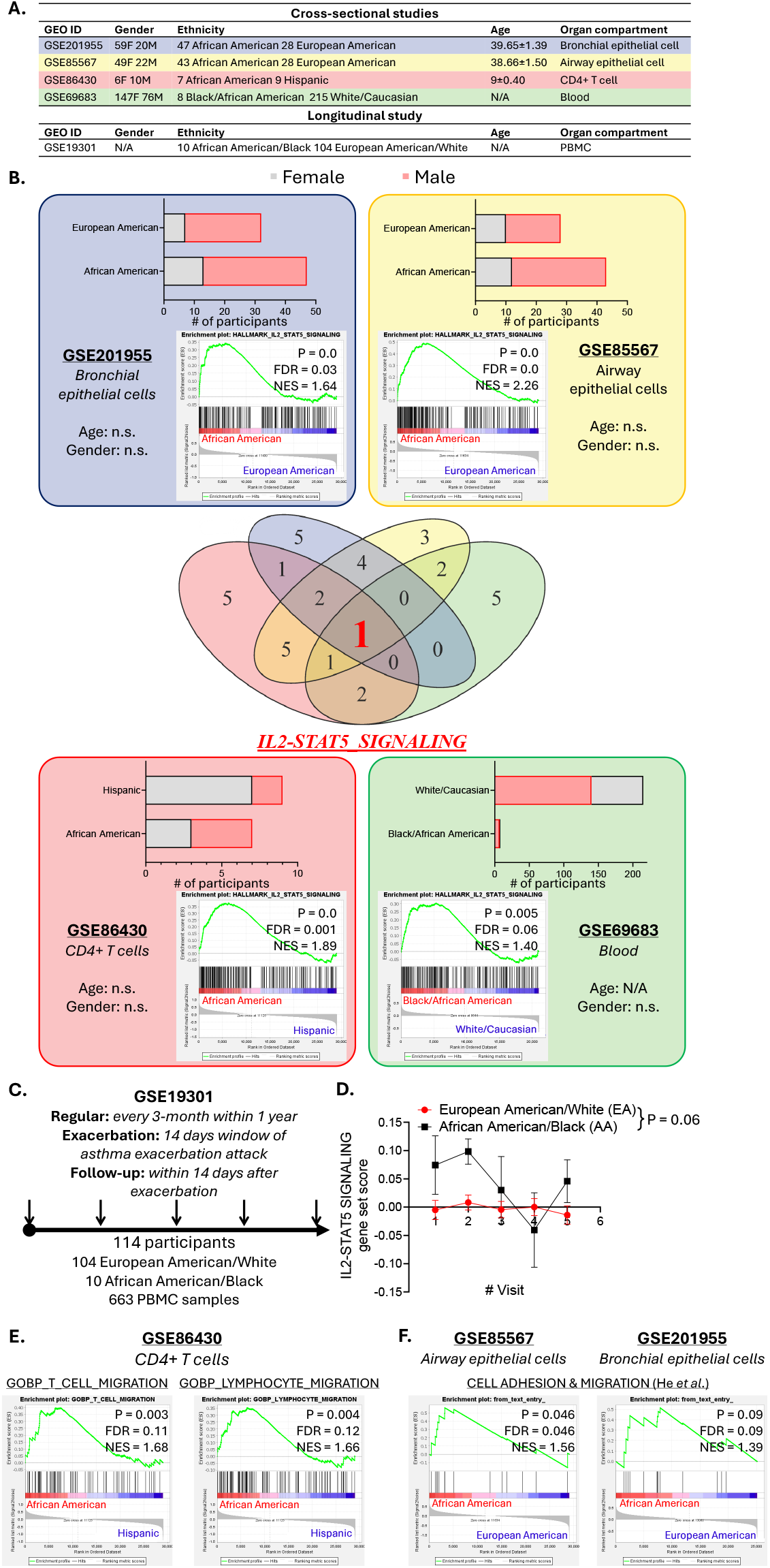
**A**. An overview for the 4 cross-sectional and 1 longitudinal studies included in present study. **B**. Gene set enrichment analysis (GSEA) of transcriptomic datasets of bronchial (blue), airway (yellow) epithelial cells, circulating CD4^+^ T cells (red), and whole blood (green) comparing African American/Black (AA) and other ethnic groups. Bar charts on the top showing the numbers of female (grey) and male (red) participants in each study. Age and gender compositions of all studies were not significantly different between ethnic groups. Within the GSEA plots AA were shown as red on the left and other ethnicities were blue on the right. Venn diagram depicted that the IL2-STAT5 signalling was the only pathway consistently enriched in AA samples. **C**. An overview of the study timeline in GSE19301. **D**. Longitudinal analysis of IL2-STAT5 signalling gene set scores in AA and European American/White (EA) peripheral blood mononuclear cells. **E**. GSEA results for gene sets related to T cell (left) and lymphocyte (right) migration comparing AA (red) and Hispanic (blue) CD4^+^ T cells. **F**. GSEA results for the gene set related to cell adhesion & migration reported by He *et al*. in airway (left) and bronchial (right) epithelial cells comparing AA (red) and EA (blue). (P: p value; FDR: false discovery rate; NES: normalized enrichment score)

The age and gender profiles for the 4 cross-sectional studies were comparably balanced (Figure2). They covered 4 different compartments (bronchial and airway epithelium, circulating CD4^+^ T cells and whole blood). In all 4 asthma datasets, gene set enrichment analysis (GSEA) revealed that AA group consistently exhibited enhanced IL2-STAT5 signalling relative to other ethnic groups (Figure2B).

The longitudinal study covering 104 EA, and 10 AA asthma patients over a period of 12 months were next analyzed. During this period, patients had regular visits every three months and/or visits for acute asthma exacerbation and a follow-up 2 weeks thereafter (Figure2C). For consistent comparison, we only focused on the routine trimonthly visits. Analysis of a total of 663 PBMC samples’ IL2-STAT5 gene set scores calculated by gene set variation analysis (GSVA) found that, longitudinally, AA generally upregulated IL2-STAT5 signalling relative to EA (Figure2D), supporting the results found in other compartments.

Mechanistically, an important contribution of the IL2-STAT5 axis to asthma development is promoting T cell migration^4^. Indeed, CD4^+^ T cells from AA showed enrichment in gene sets related to T cell and lymphocyte migration relative to the ones from Hispanic individuals (Figure2E). Likewise, AA-derived airway and bronchial epithelium exhibited elevated signalling for cell adhesion and migration implicated in asthma as previously found^4^ (Figure2F).

Together, these data illustrated that in asthma, across various compartments, African American/black seemed to upregulate IL2-STAT5 signalling, promoting T cell recruitment and migration, possibly predisposing them to more severe asthma exacerbation.

Our analyses might be subject to various confounders as asthma pathogenesis and treatment are influenced by differences in genetic backgrounds as well as various socioeconomic factors^6^. Specifically, AA patients might be more likely to be exposed to disadvantaged socioeconomic environments, predisposing them to developing asthma and have reduced access to advanced asthma treatment regimes. Hence, despite the significant differences in asthma-related signalling under both basic and disease conditions we found, future studies controlling for possible confounders with larger AA cohorts are warranted.

Collectively, our analyses for the first time decoded the in-depth mechanistic potentially underlying ethnic difference in asthma. If further validated, it will be instrumental to future asthma research and clinical practice, such as therapeutic development and stratified treatment options, taking ethnic diversity into considerations.

## Data Availability

All data produced in the present study are available upon reasonable request to the authors

## Author contributions

*Concept and design:* Duan Ni, and Ralph Nanan

*Acquisition, analysis and interpretation of data:* Duan Ni, and Ralph Nanan

*Drafting of the manuscript:* Duan Ni, and Ralph Nanan

*Critical revision of the manuscript for important intellectual content:* All authors

## Acknowledgements

This project is supported by the Norman Ernest Bequest Fund.

## Conflict of interest

Non reported.

## Notes

### Competing Interest Statement

The authors have declared no competing interest.

## References

1. Nyenhuis SM, Krishnan JA, Berry A, et al. Race is associated with differences in airway inflammation in patients with asthma. J Allergy Clin Immunol. 2017;140(1):257–257 e211.

2. Daya M, Barnes KC. African American ancestry contribution to asthma and atopic dermatitis. Ann Allergy Asthma Immunol. 2019;122(5):456–456.

3. Whitrow MJ, Harding S. Asthma in Black African, Black Caribbean and South Asian adolescents in the MRC DASH study: a cross sectional analysis. BMC Pediatr. 2010;10:18.

4. He K, Xiao H, MacDonald WA, et al. Spatial microniches of IL-2 combine with IL-10 to drive lung migratory T(H)2 cells in response to inhaled allergen. Nat Immunol. 2024;25(11):2124–2124.

5. Hondowicz BD, An D, Schenkel JM, et al. Interleukin-2-Dependent Allergen-Specific Tissue-Resident Memory Cells Drive Asthma. Immunity. 2016;44(1):155–166.

6. Ni D, Senior AM, Raubenheimer D, Simpson SJ, Macia L, Nanan R. Global associations of macronutrient supply and asthma disease burden. Allergy. 2024;79(7):1989–1989.

