## Supplementary Information for "Decoding the biology of ethnic differences in asthma"

### Materials and Methods

#### Data curation

The cellular indexing of transcriptomes and epitopes sequencing (CITE-seq) dataset analyzing healthy European American/White (EA) and African American/Black (AA) individuals was downloaded from the original publication[1] (GEO ID: GSE189050). For transcriptomic datasets, 8 asthma-related studies with ethnicity/race-related information were found on Gene Expression Omnibus (GEO, Table S1). 2 studies did not report clear ethnicity/race classification and 1 study only contained 1 AA sample. They are thus excluded. The remaining 5 datasets of high data quality and good sample ethnicity/race coverage were subject to analyses using their corresponding normalized data. For the longitudinal dataset, we only focused on the regular visit timepoints for their better data coverage and more consistent comparison as samples from exacerbation and follow-up timepoints were sporadic and analysis might be affected by bias or outliers.

**Table S1.** Overview of the asthma transcriptomic datasets used in current study.

| **Cross-sectional studies** | | | | | |
|---|---|---|---|---|---|
| **GEO ID** | **Gender** | **Ethnicity** | **Age** | **Organ compartment** | **Ref** |
| GSE201955 | 59F<br>20M | 47 African American<br>28 European American | 39.65±1.39 | Bronchial epithelial cell | 2 |
| GSE85567 | 49F<br>22M | 43 African American<br>28 European American | 38.66±1.50 | Airway epithelial cell | 3 |
| GSE86430 | 6F 10M | 7 African American<br>9 Hispanic | 9±0.40 | $CD4^+$ T cell | 4 |
| GSE69683 | 147F<br>76M | 8 Black/African American<br>215 White/Caucasian | N/A | Blood | 5 |
| **Longitudinal study** | | | | | |
| **GEO ID** | **Gender** | **Ethnicity** | **Age** | **Organ compartment** | **Ref** |
| GSE19301 | N/A | 10 African American/Black<br>104 European American/ White | N/A | PBMC | 6 |
| **Studies excluded** | | | | | |
| GSE65204 | Unclear ethnicity/race classification | | | | |
| GSE109455 | Unclear ethnicity/race classification | | | | |
| GSE172368 | Only 1 African American sample | | | | |

**Bioinformatic analysis**

CITE-seq data were analyzed with *Seurat* v4.3.0.[7]. In brief, PBMC subsets were annotated by the original publication and gene set scores were calculated using the *CellCycleScoring* function in *Seurat* based on the "IL2_STAT5_SIGNALING" gene set documented in Gene Set Enrichment Analysis (GSEA, https://www.gsea-msigdb.org/gsea/msigdb/human/search.jsp)[8]. Transcriptomic datasets were analyzed with GSEA[8] following its tutorial. For the analysis for "CELL_ADHESION & MIGRATION" gene set, the corresponding genes were adapted from the original publication[9]. Gene set variation analysis (GSVA)[10] was run following its tutorial based on the "IL2_STAT5_SIGNALING" gene set.

### References


1. Slight-Webb S, Thomas K, Smith M, et al. Ancestry-based differences in the immune phenotype are associated with lupus activity. *JCI Insight.* 2023;8(16).
2. Magnaye KM, Clay SM, Nicodemus-Johnson J, et al. DNA methylation signatures in airway cells from adult children of asthmatic mothers reflect subtypes of severe asthma. *Proc Natl Acad Sci U S A.* 2022;119(24):e2116467119.
3. Nicodemus-Johnson J, Myers RA, Sakabe NJ, et al. DNA methylation in lung cells is associated with asthma endotypes and genetic risk. *JCI Insight.* 2016;1(20):e90151.
4. Rastogi D, Nico J, Johnston AD, et al. CDC42-related genes are upregulated in helper T cells from obese asthmatic children. *J Allergy Clin Immunol.* 2018;141(2):539-548 e537.
5. Bigler J, Boedigheimer M, Schofield JPR, et al. A Severe Asthma Disease Signature from Gene Expression Profiling of Peripheral Blood from U-BIOPRED Cohorts. *Am J Respir Crit Care Med.* 2017;195(10):1311-1320.
6. Bjornsdottir US, Holgate ST, Reddy PS, et al. Pathways activated during human asthma exacerbation as revealed by gene expression patterns in blood. *PLoS One.* 2011;6(7):e21902.
7. Hao Y, Stuart T, Kowalski MH, et al. Dictionary learning for integrative, multimodal and scalable single-cell analysis. *Nat Biotechnol.* 2024;42(2):293-304.
8. Subramanian A, Tamayo P, Mootha VK, et al. Gene set enrichment analysis: a knowledge-based approach for interpreting genome-wide expression profiles. *Proc Natl Acad Sci U S A.* 2005;102(43):15545-15550.
9. He K, Xiao H, MacDonald WA, et al. Spatial microniches of IL-2 combine with IL-10 to drive lung migratory T(H)2 cells in response to inhaled allergen. *Nat Immunol.* 2024;25(11):2124-2139.
10. Hanzelmann S, Castelo R, Guinney J. GSVA: gene set variation analysis for microarray and RNA-seq data. *BMC Bioinformatics.* 2013;14:7.
